# Blind Uneven Proliferation of CD4+ T cells During Primary Infection Generates the Majority of the HIV Reservoir

**DOI:** 10.1101/2020.04.06.20053231

**Authors:** Florencia A. T. Boshier, Daniel B. Reeves, Elizabeth R. Duke, David A. Swan, Martin Prlic, E. Fabian Cardozo-Ojeda, Joshua T. Schiffer

## Abstract

The HIV reservoir is a population of 1-10 million anatomically dispersed, latently infected memory CD4+ T cells in which an HIV DNA molecule is quiescently integrated into human chromosomal DNA. When antiretroviral therapy (ART) is stopped and HIV replication initiates in one of these cells, systemic viral spread resumes, rekindling progression to AIDS. Therefore, HIV latency prevents cure. The HIV reservoir contains clones: identical HIV sequences that are integrated within identical human chromosomal DNA locations. The presence of these clones demonstrates that proliferation of CD4+ T cells sustains infection despite ART. The reservoir has a precise structure consisting of a small number of large clones and a large number of small clones. However, the mechanisms leading to this structure have not been identified. We developed a mathematical model that recapitulates the profound depletion and brisk recovery of CD4+ T cells, reservoir creation, and viral load trajectory during primary HIV infection. We extended the model to simulate stochastically individual HIV reservoir clones and identified that uneven proliferation among clones during recovery from CD4+ lymphopaenia is sufficient to explain the observed clonal reservoir distribution. We project that within one month of infection 75-95% of reservoir cells are generated from cellular proliferation rather than denovo viral infection. Recent detection of HIV infected clones during the first 5 weeks of infection support our model’s predictions.

---

The HIV latent reservoir is a population of resting memory CD4+ T cells that contain HIV-1 DNA integrated within their human chromosomal DNA [40, 27, 26, 16]. Resumption of HIV replication within one latently infected cell can spark systemic spread of virus, usually within days to weeks of antiretroviral therapy (ART) analytical treatment interruption (ATI) [32, 20, 39, 78]. The HIV reservoir size is stable over decades of ART [70, 23] and imperturbable, even if pharmacologically powerful agents are given that reactivate the virus in cell culture [69, 8]. Persistence of HIV latency therefore precludes cure and necessitates lifelong ART.

The HIV reservoir is established swiftly after initial infection. In non-human primate models, reservoir seeding occurs within three days of viral challenge [77, 76]. In humans, initiation of ART within 10 days after estimated HIV infection leads to a reduced latent reservoir size [6] and substantial delays in viral rebound after ATI [41, 30, 28, 22] suggesting that the reservoir formation begins in the first week after HIV acquisition [31, 5].

Long-term studies of individuals on prolonged ART show that the reservoir often contains multiple cells carrying HIV DNA proviruses with identical sequences [36, 18, 48, 54, 15, 73, 72], which are integrated into the exact same human chromosomal integration site [73, 45, 25, 19]. Each unique sequence is believed to be generated from a distinct cellular infection event, as the HIV reverse transcriptase enzyme is highly error prone [79]. Identical viral sequences with matching integration sites suggest that these genomes were copied by the cellular DNA polymerase during the S phase of interphase, prior to mitosis. Consequently, the existence of abundant HIV DNA sequence clones proves that proliferation of infected memory CD4+ T cells helps sustain the HIV reservoir [59].

The clonal structure of the HIV reservoir appears to be highly organised. The rank according to size of an individual HIV cellular clone is likely to be predictive of the size of that clone. The reservoir is in turn defined by a small number of high-rank, large clones and a far larger number of low-rank, small clones [59]. This trend is evident when all HIV DNA is measured or when replication competent viruses are assessed alone [37]. A similar structure is observed for clones within the entire T cell repertoire as defined by T cell receptor (TCR) sequencing [42, 24, 55], which suggests that HIV reservoir clone dynamics may be partially or fully governed by more general rules of CD4+ T cell immunity [67]. However, the exact mechanisms and cellular dynamics leading to the precise rank-order distribution of the HIV reservoir are not known.

The HIV reservoir’s clonal structure is clearly evident after a year or less of ART initiation and has been observed in individuals who started ART soon after primary infection [37, 34, 45, 73], suggesting that viral and cellular dynamics during early infection contribute to its formation. Primary HIV infection is characterised by profound CD4+ T cell lymphopaenia and peak viremia two weeks post-infection, followed by an incomplete, but brisk recovery in CD4+ T cell levels over the next four weeks. Stable HIV DNA and RNA setpoints are established concurrently approximately four weeks post-infection [5, 53, 63].

Formation of the HIV reservoir may be impacted by several coupled, non-linear processes including a mounting immune response [53], CD4+ T cell activation [50], and target cell limitation [61], necessitating the use of mathematical models to decipher available data. In previous modelling studies, generation of latently infected cells during untreated primary infection was ascribed entirely to viral infection of memory CD4+ T cells with subsequent conversion to a resting state [70, 9, 32, 60]. CD4+ T cell reconstitution is likely to be driven by homeostatic proliferation. Mouse models of CD4+ T cell recovery following depletion conclude that homeostatic proliferation is blind at the cell subset level with overall competition between CD4+ and CD8+ T cells for limited resources and space during homeostatic proliferation [49, 2, 3, 64, 44] but highly uneven at the single clone level-with some TCR-defined clones outcompeting others due to variable binding affinity to self-antigens [3].

Here we develop a model of HIV primary infection that includes blind and uneven homeostatic proliferation of latently infected cells during recovery from CD4+ lymphopaenia. We fit the model to empirical data from 12 untreated individuals during primary infection [43, 52]. The model output suggests that homeostatic proliferation of CD4+ T cells drives the observed size and clonal structure of the HIV reservoir during the first several weeks of infection. It makes the empirically confirmed prediction that HIV sequence clones should be discoverable within a month of HIV infection despite the fact that most circulating HIV DNA sequences are in actively infected cells with unique non-clonal sequences [17].

## 1 Methods & Materials

### 1.1 Deterministic mathematical model

We developed several competing mathematical models to study the potential role of homeostatic proliferation in the formation of the latent reservoir during primary infection. All candidate models are derived from the following master system:

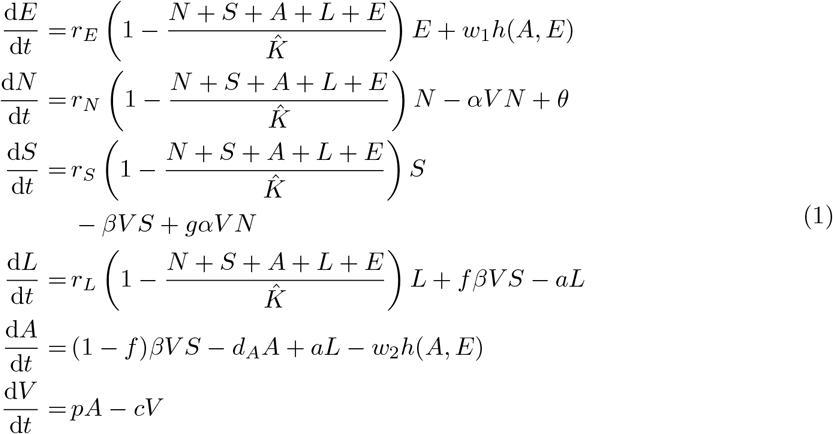

Where *E*(*t*) represents the concentration [cells/*µ*L] of CD8+ T cells at time *t*. The CD4+ T cells are divided into four compartments: non-susceptible, susceptible, latently infected and actively infected. We denote the concentration [cells/*µ*L] of these at time *t* by *N* (*t*), *S*(*t*), *L*(*t*) and *A*(*t*) respectively. For data fitting purposes, the reservoir of latently infected cells *L*(*t*) represents the total HIV DNA rather than the much smaller subset of genomically intact or replication competent sequences [34]. Finally, *V* (*t*) represents the plasma viral load over time [copies/*µ*L]. We use the term all infected cells to refer to the sum of actively and latently infected cells.

The viral component of the model builds upon the canonical model of viral dynamics by differentiating activated and non-activated CD4+ T cells [33, 62, 75]. The virus is cleared at a rate *c*. Actively infected cells produce virus at a rate *p*, which describes the aggregate rate of constant viral leakage and burst upon cell death, and die at a rate *d*_*A*_. The susceptible cells, *S*, are infected by the virus at a rate *β* [*µ*L/virus -day]. We assume that a fraction, *f*, of infection events generate latently infected cells and the remaining fraction, (1 − *f*), leads to productively infected cells. Latently infected cells reactivate and become productively infected at a rate *a* [cell/*µ*L - day].

In contrast to previous infection models, we incorporate the blind homeostasis hypothesis to explain cellular dynamics. We assume that there is an inherent competition for resources between CD8+ and CD4+ T cells that impact expansion and cell survival. We model this by logistically limiting the growth of the CD8+ T cells and the non-actively infected CD4+ T cell subsets by their sum. A similar mathematical formulation of blind homeostasis was considered in [49] to explain experiments of T cell reconstitution following CD4+ depletion in mice [3]. The turnover rates [cells/*µ*L - day] are denoted by *r*_∗_. The functional carrying capacity of the system is denoted by *K* [cells/*µ*L]. We considered a set of models that did not include this assumption by excluding the CD8+ T cell compartment entirely.

We evaluated the importance of several additional mechanisms. First, the killing of actively infected cells by CD8+ T cells at a rate *w*_2_, with or without the HIV antigen stimulation of CD8+ T cells at a rate *w*_1_ as described by function *h*(*A, E*). We consider two forms for *h*(*A, E*), either linear or concentration-dependent as given by equation (2).

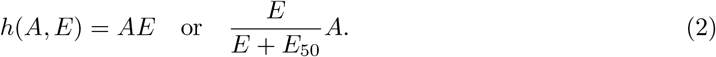

Second, we considered the possibility that the virus removes non-susceptible cells from their compartment at a rate *α* [*µ*L/virus -day]. A proportion (1 − *g*) undergo bystander killing [47] and the remaining fraction become susceptible, i.e. upregulate CCR5, as they are part of the mounting cellular immune response to the virus. Finally, we investigated the possible role of generation of non-susceptible cells by the thymus at a constant rate *θ*. We assessed the above assumptions individually and in combination. Overall, we constructed 12 competing models. A detailed account of the model selection can be found in *Supporting Information S1*.

### 1.2 Stochastic mathematical model of individual clones within the latent reservoir

We are principally concerned with whether and how the proliferation of latently infected cells affects formation and sustainment of the clonal structure of the HIV reservoir during primary infection. To study the clonal distribution of the latent reservoir, we use a stochastic model in-place of equation 7. We let 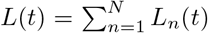, where *L*_*n*_(*t*) denotes the number of cell members in clone *n* at time *t* and *N* denotes the number of distinct clones. In any short time-interval, four events can occur: a new distinct sequence can be introduced into the latent reservoir, an existing member of a clone can proliferate, a clone can contract in size or extinguish, or an existing member of a clone can convert to an actively infected cell and reactivate HIV.

Each new viral infection of a cell is associated with a unique HIV DNA sequence due to the error prone HIV reverse transcriptase, as well as a unique human chromosomal integration site [45, 54, 65, 66]. Therefore we assume that each infection event that generates a latently infected cell introduces a new viral sequence. We assume each new sequence population to have an initial size of 1, such that

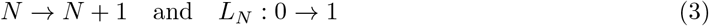

with probability *fβV* (*t*)*S*(*t*).

Each clone in the reservoir may die out stochastically or expand due to cellular proliferation. We refer to a clone with a single member, *L*_*n*_ = 1, as a singleton. Upon proliferation, a single HIV DNA molecule is copied with the human DNA polymerase, generating equivalent HIV DNA sequences and human chromosomal integration sites in the two daughter cells. Therefore we allow each clone in the reservoir to grow (*L*_*n*_ → *L*_*n*_ + 1) with probability *r*_*L,n*_*L*_*n*_*/K* and decline (*L*_*n*_ → *L*_*n*_ − 1) with probability (*r*_*L,n*_(*N* + *S* + *I* + *L* + *E*)*/K* + *a*)*L*_*n*_. When *L*_*n*_ *>* 1 its HIV DNA sequence is considered to be part of a true clone in our model output.

The parameter *r*_*L,n*_ represents the proliferation rate of the clone *n*. Based on the observations that distinct T cell clones proliferate at vastly different rates in response to lymphopaenia [3], and that the HIV reservoir is characterised by variability in clone size over 5-6 orders of magnitude [59], we allowed each clone to take on a different proliferation rate. Specifically, we defined uneven proliferation by assuming that the turnover rates are lognormally distributed, *r*_*L,n*_∼*lognormal*(*µ, σ*) with corresponding mean *m* = exp(*µ* + *σ*^2^) and variance *v* = (exp(*σ*^2^) −1) exp(2*µ* + *σ*^2^).

Our simulations were run in Matlab. We simulated the deterministic compartments using ode45 [68] and used a Gillespie tau-leap of the same time-step for the stochastic compartment [29]. Each was updated simultaneously. Simulations were run in a litre of blood to simulate observed clinical values and to avoid numerical intractability of whole body simulations.

### 1.3 Parameter estimation

We fit our deterministic primary infection model (instances of equation (1)) to digitally extracted viral loads and matching CD4+ counts from 12 participants from the Females Rising through Education, Support and Health cohort (FRESH) [43, 52]. Our goal was to capture generalised trends in viral expansion, peak, contraction and setpoint, and CD4+ T cell contraction, re-expansion and re-establishment of a setpoint. We employed a nonlinear, mixed-effects modelling approach implemented in Monolix [7]. Full details of model fitting can be found in *Supplementary information S1*. We fixed 9 of 17 parameters from reported values in the literature and estimated the remaining 8. For fitting the deterministic model we made a simplifying assumption that *r*_*L*_ = *r*_*N*_.

We also fixed the initial conditions of the different variables in our model. We set *A*(0) = 1 and *L*(0) = *V* (0) = 0. As we have no CD8+ T cell data, we set *E*(0) = 500, the mean pre-infection values reported in the RV217 study [63], for models requiring it. We assume that susceptible cells are represented primarily by CCR5+ T cells, which in this cohort are found to be between 1-12 % of total CD4+ T cells in the plasma pre-infection [52]. We therefore set *S*(0) to be 10% and *N* (0) to be 90% of total CD4+ T cells for all participants. The resulting carrying capacity is found by assuming steady state pre-infection such that *K* = *E*(0) + *S*(0) + *N* (0).

The parameters *µ* and *σ* governing clone proliferation rates and the latency fraction *f* are unknown. Therefore we performed sensitivity analyses to examine the effects of these three parameters on the size of the latent reservoir throughout infection and its clonal structure at 50 days post-infection.

## 2 Results

### 2.1 Blind proliferation between T cells explain CD4+ reconstitution during primary infection

The most parsimonious model by Akaike Information Criterion (AIC) score is shown in figure 1 and mathematically here:

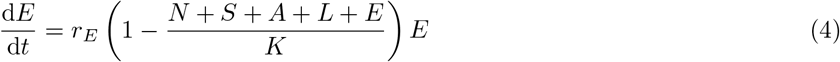

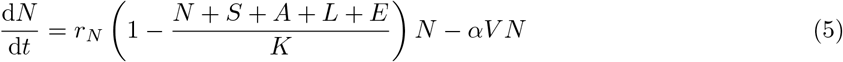

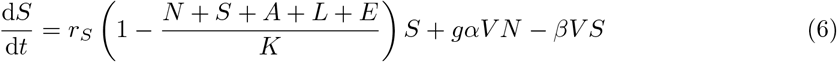

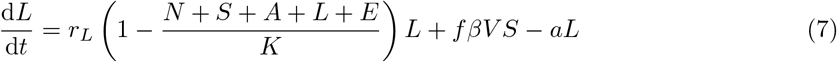

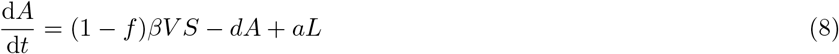

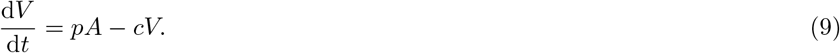

**Figure 1:**
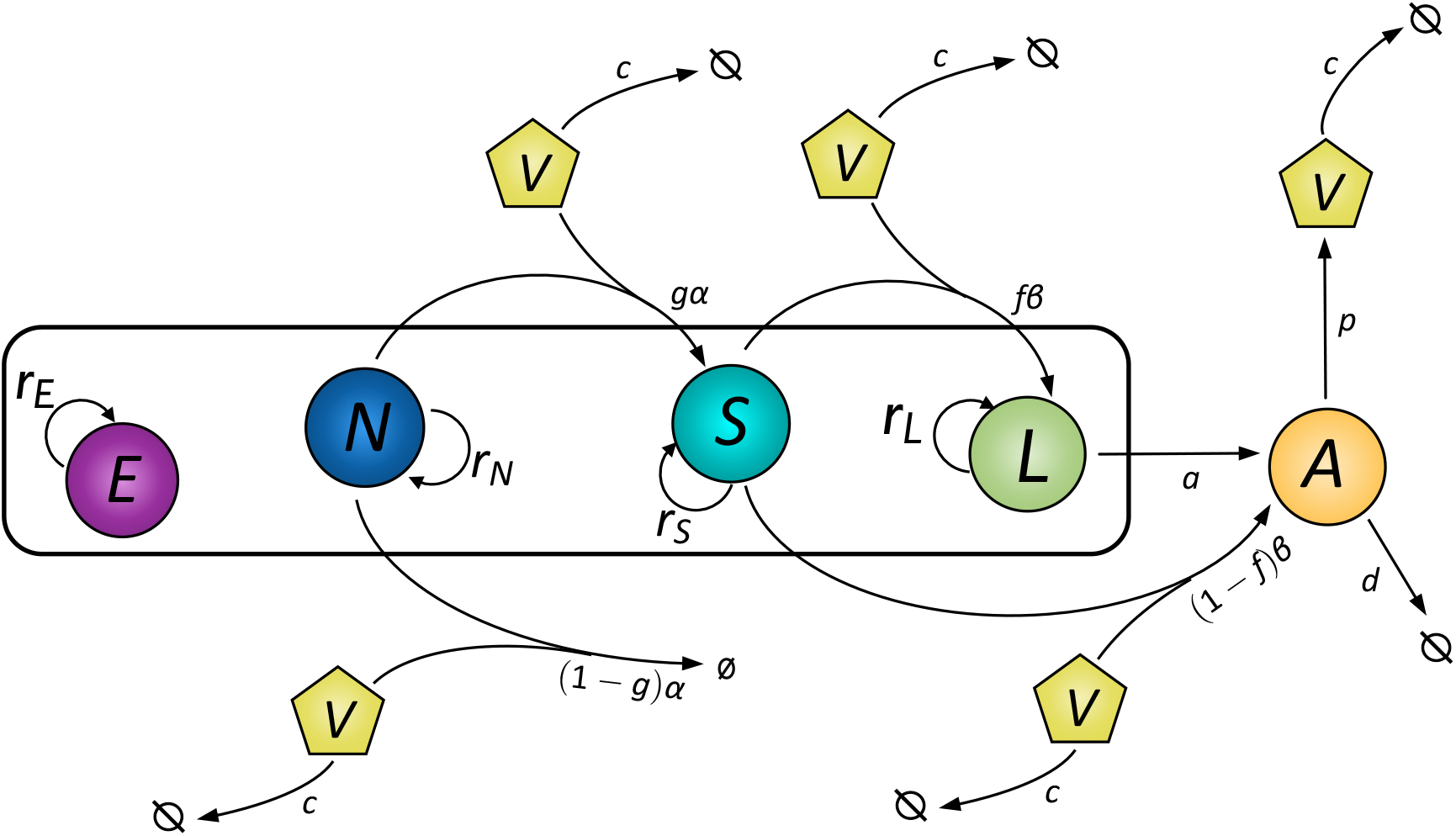
Most parsimonious mathematical model of primary HIV infection. *E* represents CD8+ T cells. The CD4+ T cells are divided into four compartments: non-susceptible (*N*), susceptible (*S*), latently infected (*L*) and actively infected (*A*). The virus is represented by *V*.

Key assumptions required for model fit to data include blind homeostatic proliferation of both CD4+ and CD8+ T cells, density-dependent virus induced bystander killing of non-susceptible CD4+ T cells, and density-dependent virus induced activation of non-susceptible CD4+ T cells to susceptible CD4+ T cells via upregulation of CCR5+. CD8+ T cell expansion is driven solely by blind homeostatic proliferation. As studies in humans with hyperacute infection have shown that 5-20 % of activated CD8+ T cells during primary infection are responding to the virus [52] we also simulated an alternative model that incorporates HIV-induced CD8+ T cell activation. This model also fits the data but had a limited impact on latent reservoir dynamics as shown in figure S1.

The population level parameters found under these assumptions are shown in table 1. Certain solved model parameters correlate, hence we do not claim to have identified their absolute values. The individual parameter values used for each participant can be found in Supplementary table S2. In figure 2, we show the model fit to data from a single study participant. For measurements of cellular concentrations, we scale simulation results, which are run in a liter of blood, to a *µL* and state viral load results in *mL*. Simulation results from our model closely recapitulate longitudinal measures of viral load (figure 2a) and total CD4+ counts, which is the sum of *N* (*t*), *S*(*t*), *L*(*t*) and *A*(*t*), (figure 2b). The expansion behaviour of the CD8+ T cell trajectories (figure 2c) while not available to us from this study, falls within the range observed during primary infection in other studies [63]. The reservoir volume (measured as total reservoir HIV DNA in figure 2d) expands to levels observed during chronic ART treatment [14] with an HIV DNA setpoint within 2 weeks of infection. Model fits for the other study participants are shown in figure S1. Predicted trajectories for *N* (*t*), *S*(*t*), *L*(*t*) and *A*(*t*) suggest that each of these cell populations equilibrate to a quasi-steady state 20-40 days after infection.

**Table 1:** Summary of deterministic model parameters and initial conditions. When estimated, the value shown is from the population level fits

| Parameter | Value | Dimension | Source |
| --- | --- | --- | --- |
| $t_0$ | -3.76 | days | estimated |
| $E_0$ | 500 | cells/μL | [63] |
| $N_0$ | $0.90 \times T_0$ | cells/μL | fixed |
| $S_0$ | $0.10 \times T_0$ | cells/μL | fixed |
| $K$ | $E_0 + N_0 + S_0$ | cells/μL | fixed |
| $r_E$ | 0.085 | cells/(μL-day) | estimated |
| $r_N$ | 0.251 | cells/(μL-day) | estimated |
| $r_S$ | 0.311 | cells/(μL-day) | estimated |
| $r_L$ | 0.251 | cells/(μL-day) | assumed equal to $r_N$ |
| $\alpha$ | $1.2 \times 10^{-5}$ | μL/(virus-day) | estimated |
| $g$ | 0.26 | NA | estimated |
| $f$ | $10^{-4}$ | NA | [21] |
| $\beta$ | $10^{-4.01}$ | μL/(virus-day) | estimated |
| $d$ | 1 | day$^{-1}$ | [46] |
| $p$ | $10^4$ | virus/(cell-day) | [56, 35] |
| $c$ | 23 | day$^{-1}$ | [58] |
| $a$ | $5.75 \times 10^{-5}$ | day$^{-1}$ | [4] |

**Figure 2:**
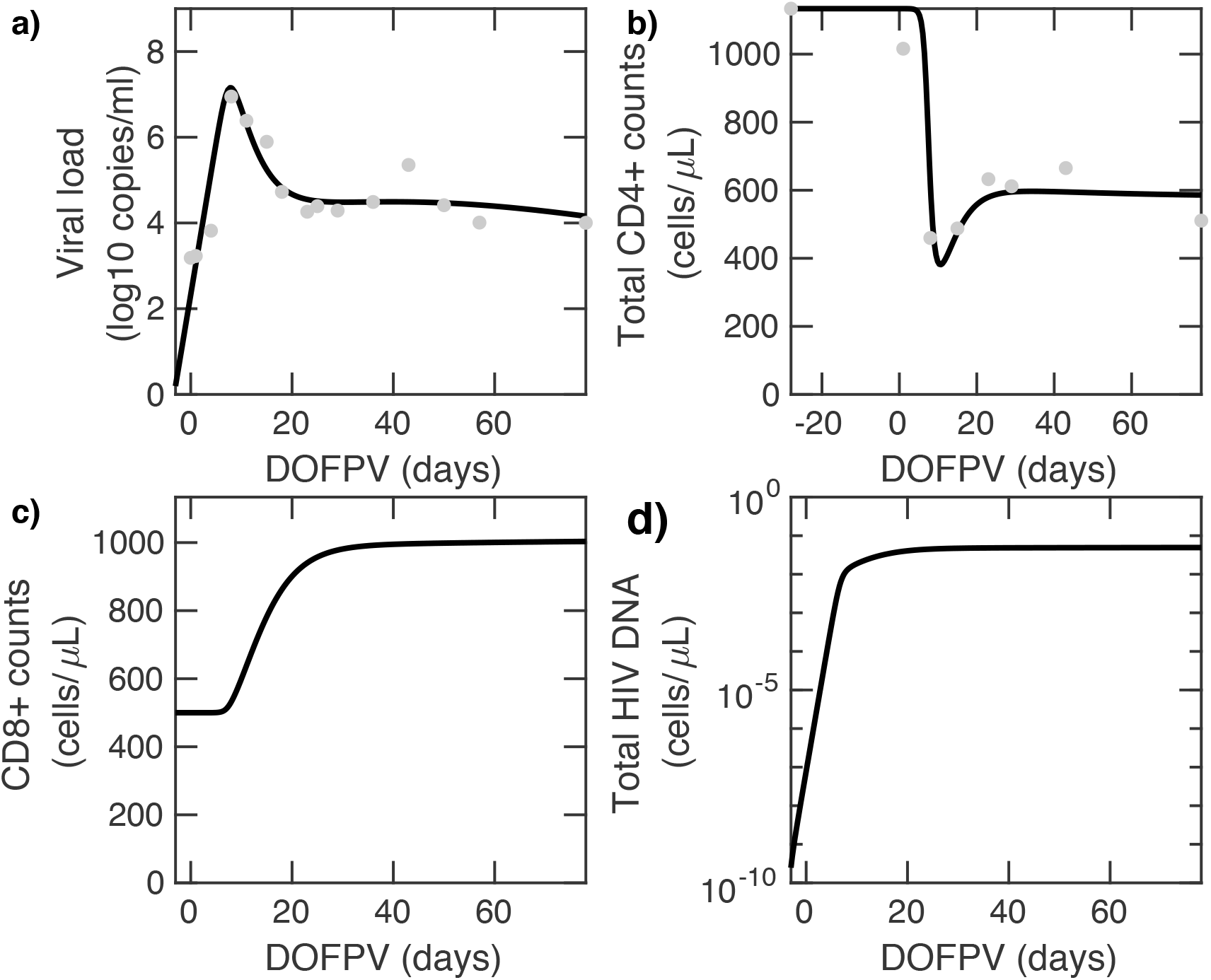
Longitudinal results of deterministic mathematical model fits (black line) to data from participant 11 (dots) for: a) viral load b) CD4+ T cells = *N* (*t*) + *S*(*t*) + *A*(*t*) + *L*(*t*), c) CD8+ T cells d) Latent reservoir size. X-axis shown in days following onset plasma viraemia (DOFPV).

### 2.2 Homeostatic cellular proliferation is required to recapitulate the inverse correlation observed between CD4+ T cell nadir and HIV reservoir volume

Previous studies identified that levels of HIV-1 proviral DNA during chronic ART are negatively correlated with CD4+ T cell nadir during primary infection [11] particularly if ART is started during acute infection [71]. In order to establish whether this correlation occurs during untreated primary infection in our simulations, we compare CD4+ T cell nadir and size of the latent reservoir at 30 days post-infection under our deterministic model as shown in figure 3. Simulating all participants using parameter values derived from our individual fits, table S2, we identify that CD4 T-cell nadir as estimated from the model had a significant, negative correlation with size of the latent reservoir 30 days post-infection (*r*^2^ = 0.34, p¡0.05). At later times during infection, the correlation weakens in agreement with [71]. However no correlation exists if reservoir proliferation is excluded (i.e. set *r*_*L*_ = 0), which suggests that the observed correlation is mechanistically related to latent cell proliferation during primary infection.

**Figure 3:**
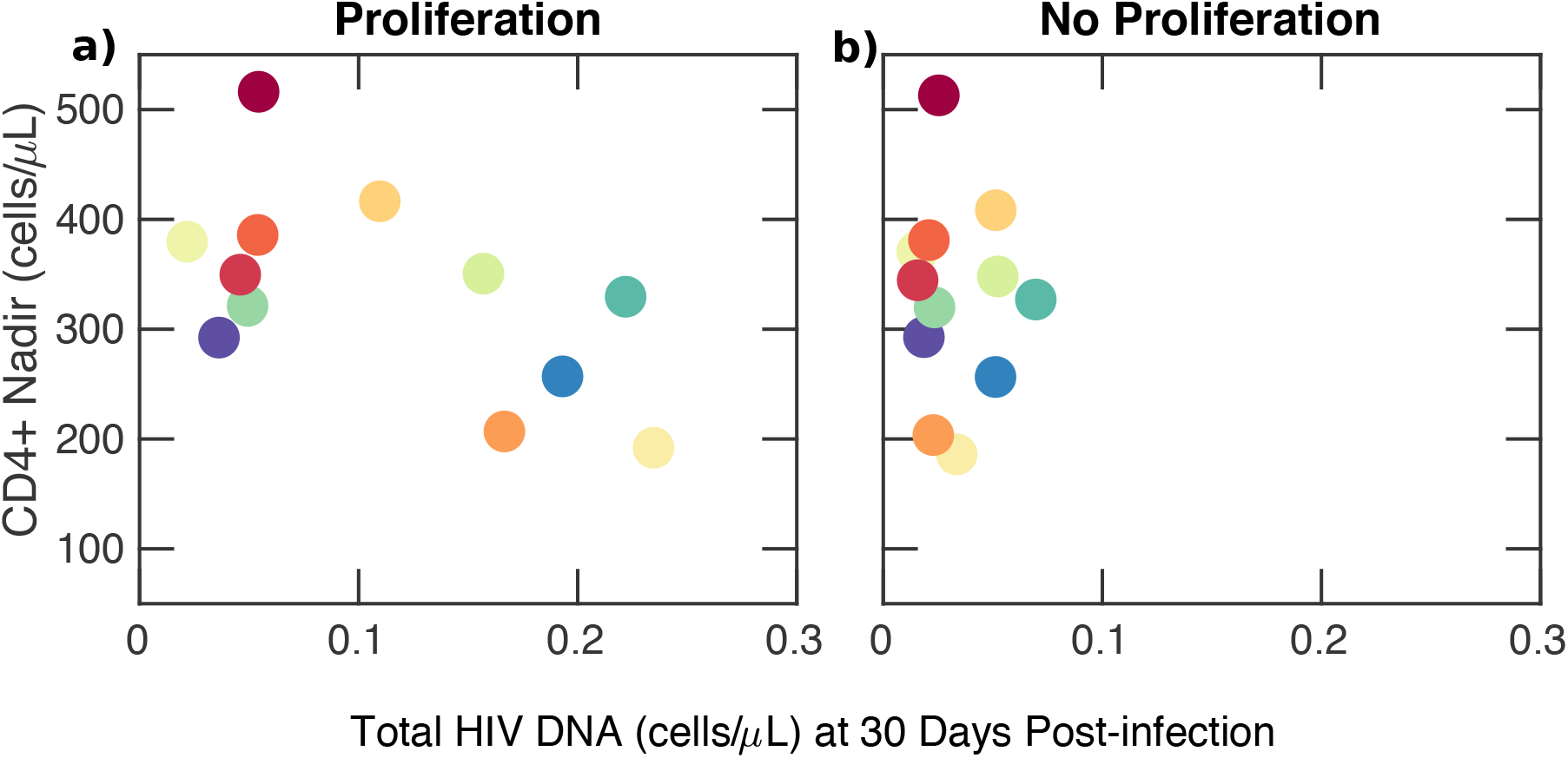
Inverse correlation between CD4+ nadir and latent reservoir volume at 30 days after primary infection in model simulations. a) An inverse correlation between the latent reservoir and CD4 + T cell nadir exists if the latent reservoir proliferates (*r*_*L*_ ≠ 0) (p¡0.05). Each colour corresponds to one of twelve participants in the FRESH cohort. b) Correlation between the latent reservoir and CD4 + T cell nadir disappears if the latent reservoir does not to proliferate (*r*_*L*_ = 0).

### 2.3 Individual clone expansion is predicted to occur immediately after reservoir formation

Next we simulated our stochastic model with individual clones. A single realization of the latent reservoir is shown in figure 4. Expanding clones are observed in the latent reservoir less than a day after it is seeded, and new HIV DNA sequences are added to the reservoir continuously throughout untreated infection due to new infection events. The individual clones sum to a reasonable approximation of reservoir size, comparable to the deterministic model simulations in figure 2d.

**Figure 4:**
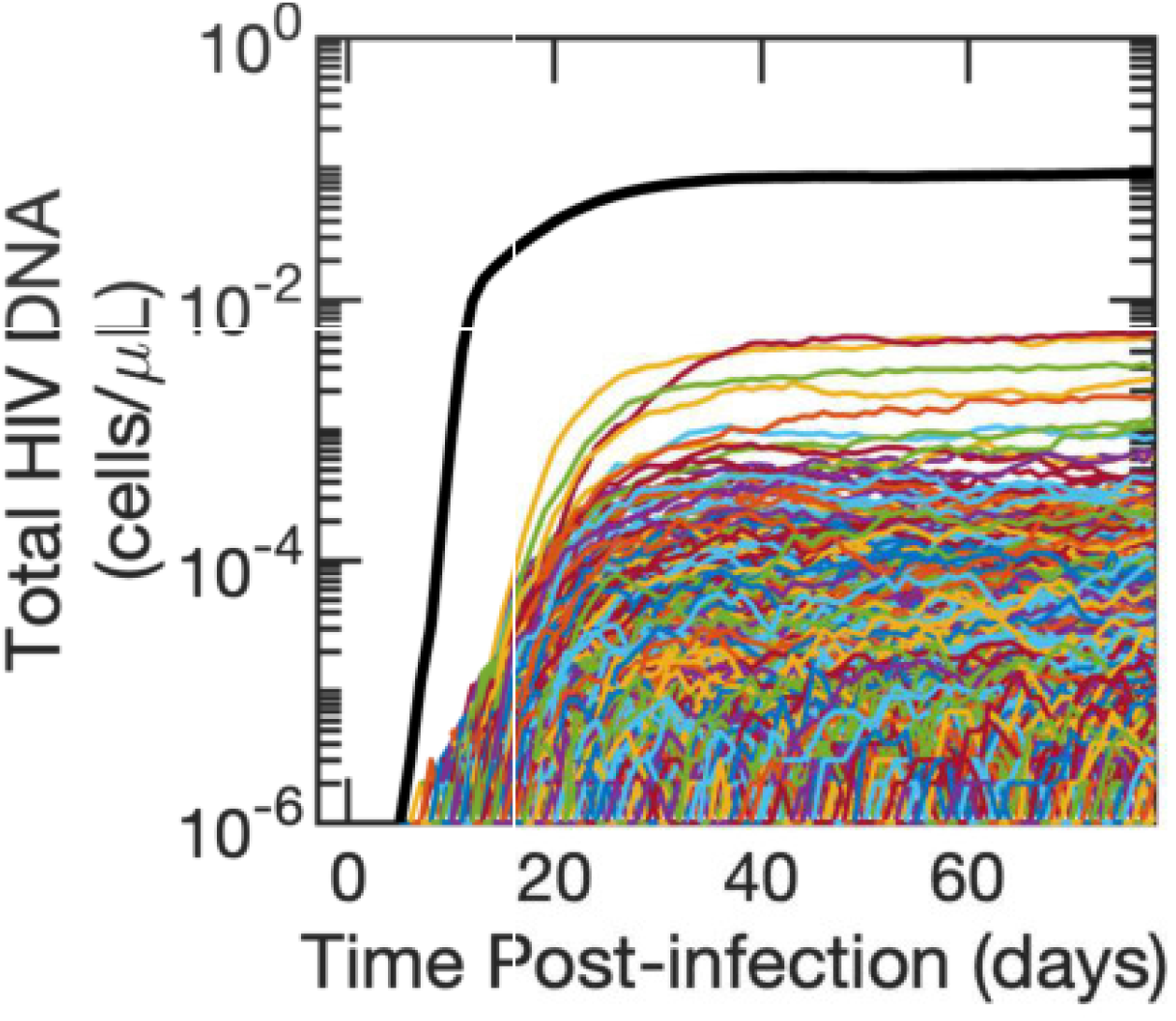
Simulated single clone dynamics during primary HIV infection. Each clone is represented by a single coloured line. The sum of all clones is shown in black. The mean of *r*_*L*_, *m* = *r*_*N*_ and the variance *v* = 0.2*r*_*N*_.

Assuming otherwise unchanged infection dynamics, HIV reservoir size could potentially be determined by three parameters in our stochastic model: the latency fraction, *f* (fraction of newly infected cell entering latency), mean clone proliferation rate, *m*, and variance of proliferation rates across clones, *v*. We find that altering the latency fraction by several orders of magnitude has a negligible impact on the projected size of the reservoir (figure S2). Increasing latent cell proliferation rates to twice those in other non-susceptible CD4 T cells results in several-fold larger reservoir volumes (figure S2a, S3a). Increasing the variance in proliferation rate across reservoir clones also generates larger reservoir volumes (figure S2b, S3b) and increases the sensitivity of the model to latency fraction in a non-linear fashion (figure S2b). Whilst moderate mean proliferation rates and variances lead to plausible latent reservoir sizes in stochastic model simulations, very large mean rates and/or variances may result in unrealistically large latent reservoirs comprising more than 50% of an individual’s CD4+ T cell repertoire (figure S3). Therefore we are able to establish upper bounds on values for the mean and variance of latent cell proliferation rate.

### 2.4 The latent reservoir reaches its highest sequence richness during the first 10 days of HIV infection

Next we consider the dynamics of sequence richness of the latent reservoir. Sequence richness is the number of unique total HIV DNA sequences or chromosomal integration sites in the body and is a marker of the number of latent cells generated by de novo HIV infection. Mathematically, we define sequence richness at time *t, R*(*t*), as

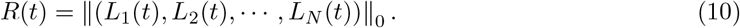

As shown in figure 5a, in model realizations of all study participants, the reservoir rapidly accumulates richness during the first∼10 days of infection due to unchecked viral replication leading to widespread infection of new cells with unique sequence and chromosomal integration sites. Following peak viraemia, the richness declines gradually towards a steady state as stochastic burnout of some clones balances formation of new sequences due to new ongoing infection events.

**Figure 5:**
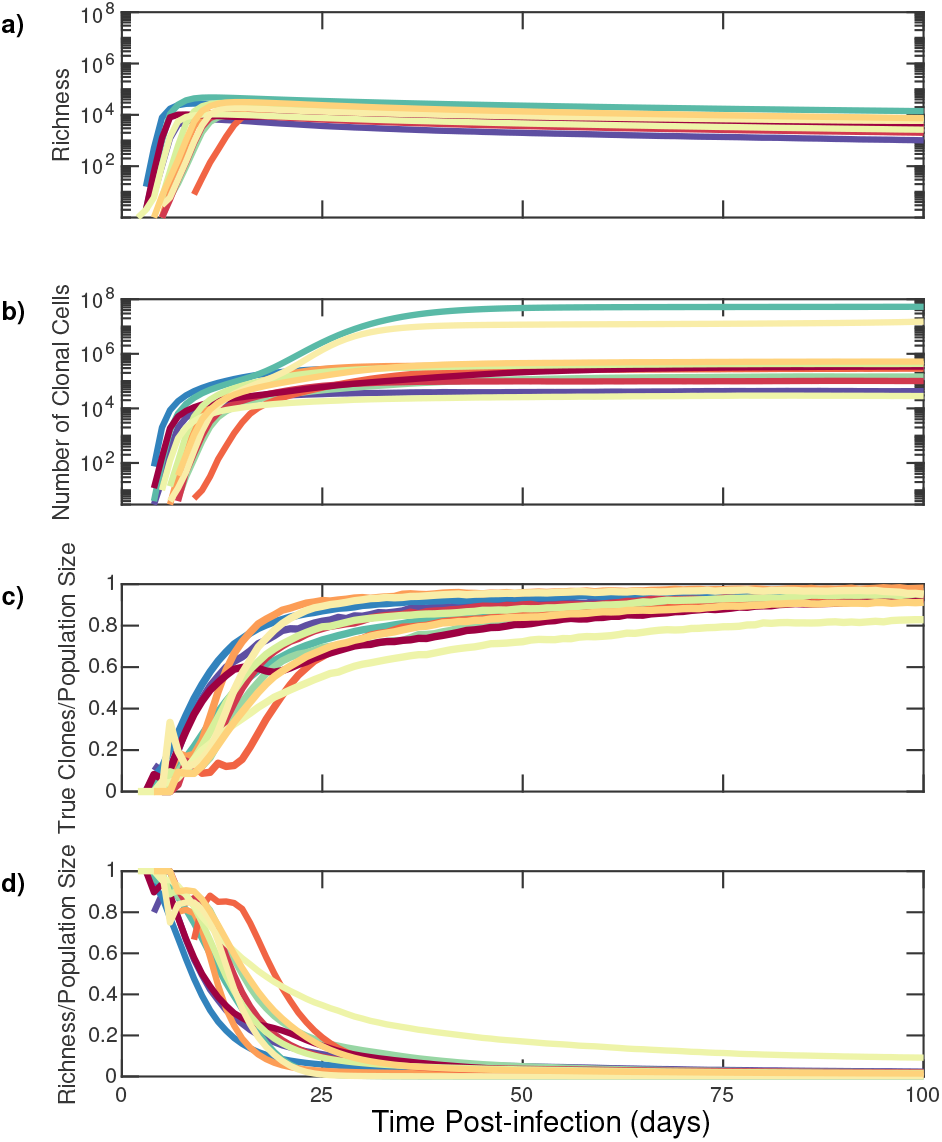
Establishment of clonal predominance in the HIV reservoir within the first month of infection due to rapid homeostatic proliferation of CD4+ T cells from stochastic model simulations. (a) Richness of HIV DNA sequences within latently infected reservoir cells. (b) Number of HIV DNA reservoir sequences within proliferative CD4+ T cell clones. (c) Clonal sequence : total sequence population size ratio for latently infected cells. (d) Richness : total sequence population size ratio for latently infected cells. Each colour corresponds to an individual as in figure 3.

### 2.5 Proliferative clones predominate in the latent reservoir within two weeks of primary HIV infection

In model simulations, expansion of sequence richness terminates at the CD4+ T cell nadir∼10 days post infection (figure 2b,d). This is followed by 2-4 weeks of rapid blind proliferation of CD4+ and CD8+ T cells to allow recovery from CD4+ lymphopaenia. As part of this process, homeostatic proliferation occurs among individual CD4+ T cell clones within the latent reservoir (figure 5b). Within two weeks of infection 12% to 71% of the latent reservoir is predicted to consist of clones derived from proliferation; within 100 days of infection 82% to 98% of the latent reservoir consists of clones derived from proliferation (figure 5c).

We normalised sequence richness according to the total number of latently infected cells (figure 5d). The maximum value of this ratio approaches 0.88-1.0 within the latent reservoir 10 days after infection as nearly all reservoir HIV DNA sequences are unique prior to clonal expansion. However, this ratio rapidly declines over the next 2 weeks to 0.5 by day 9-20 and 0.1 by day 18-88. This result implies that while viral infection establishes the reservoir extremely early after HIV transmission, the reservoir is maintained primarily by clonal cellular proliferation in response to CD4+ lymphopaenia following peak viraemia.

### 2.6 Proliferative reservoir clones are obscured by a larger population of non-clonal sequences within actively infected cells during untreated infection

There are currently no experimental methods available to isolate latent reservoir HIV DNA sequences from sequences within cells harbouring active HIV replication during untreated infection. Instead, the latent reservoir is obscured by a population of rapidly mutating sequences from actively infected cells, each with a unique genetic signature and chromosomal integration site.

However, we can distinguish latently infected cells from actively infected cells in our model. Therefore we performed the above analysis inclusive of actively infected cells, each of which contributes a unique sequence singlet to the overall population of infected cells. Early during untreated infection, the number of actively infected cells outnumbers the number of latently infected cells by 1-3 orders of magnitude, such that observed richness is driven mostly by actively infected cells (figure 6a compared to figure 5a). During the first 10 days of infection, the proportion of clones among all sequences is extremely low (range: 10^−5^ - 6 *×* 10^−4^) (figure 6b).

**Figure 6:**
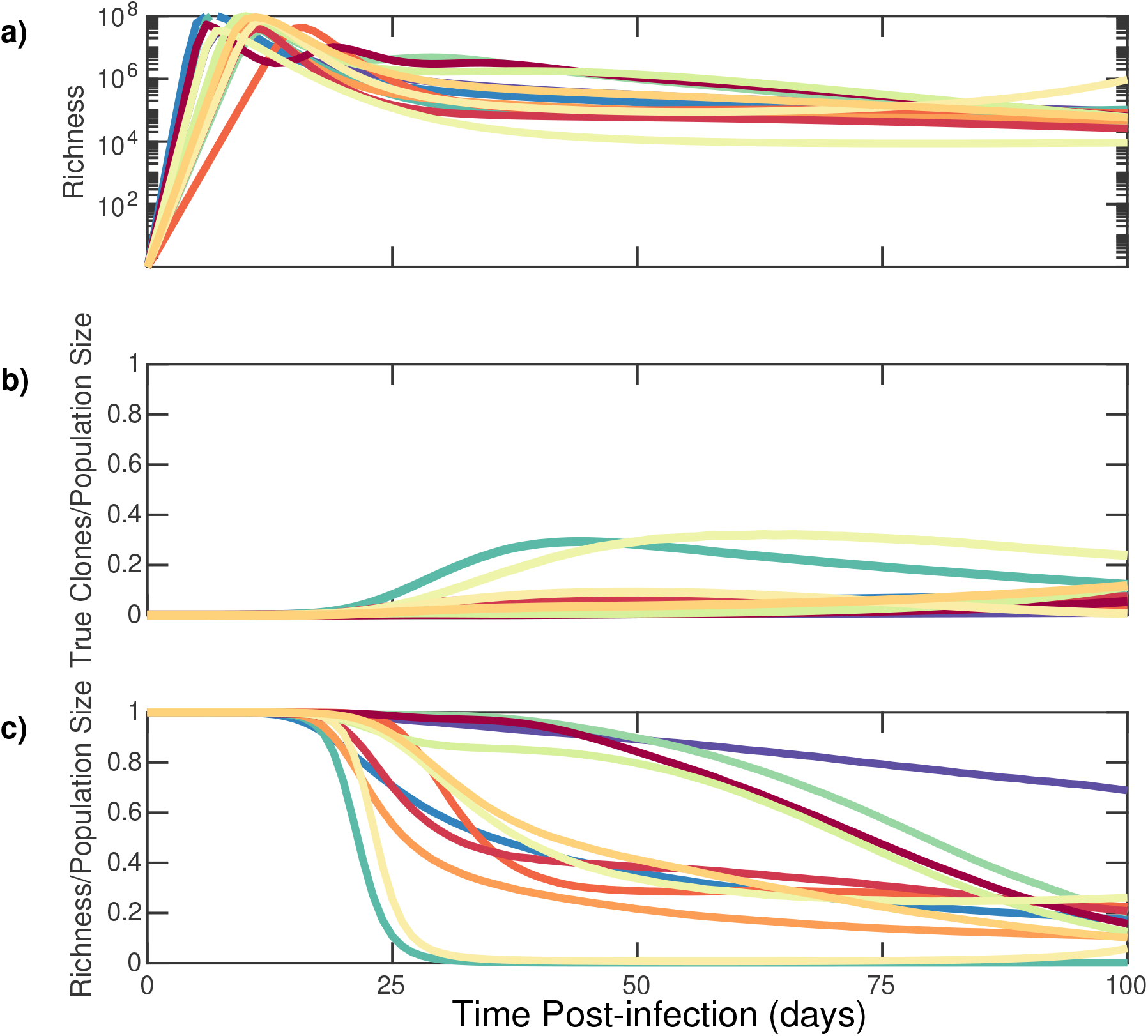
HIV DNA reservoir sequences are obscured by a cloud of non-clonal sequences from actively infected cells during early untreated infection in stochastic model simulations. (a) Richness of HIV DNA sequences within all infected cells. (b) Clonal sequence : total sequence population size ratio within all infected cells. (c) Richness : total sequence population size ratio within all infected cells. Each colour corresponds to an individual as in figure 3.

Upon establishment of a viral load and reservoir steady state at 25-50 days, most HIV DNA sequences obtained from a clinical sample are truly unique and the ratio of clones to population size is still relatively low (range: 5 × 10^−3^ - 3 ×10^−1^) figure 6b, as actively infected cells still outnumber clonally expanding latently infected cells. Accordingly, the ratio of richness to the total population remains high figure 6c. Within 100 days of infection 2 *×* 10^−3^ to 7 *×* 10^−1^ of all infected cells are predicted to consist of clones figure 6b.

We examined whether these predictions were robust to uncertainty regarding key reservoir model parameters. Lower mean proliferation and higher variance in proliferation rate results in slight delays in the timing of clonal predominance within the HIV reservoir as does higher variance in turnover rates (figure S4a,b,c,d). While these parameters may indeed vary across infected persons, it is a generalisable property of our model output that a majority of the latent reservoir (figure S4a,c), but a minority of all infected cells (figure S4b,d) are composed of sequence clones within a month of infection.

### 2.7 Adequate sequence sample size ensures detection of HIV DNA clones within 2-3 weeks of primary infection

Our model projects that by Fiebig stage IV (day 27), 70-95 % of HIV DNA sequences within the reservoir are clonal (figure 5 c). However, only 2-20% of total HIV DNA sequences are clonal (figure 6b) making the detection of clones in the reservoir challenging. Another barrier to detecting clonal HIV DNA from the reservoir is that the sequence sample size in most reservoir studies are low, varying between ∼10 in studies studying intact, replication competent viruses and ∼ 100-1000 in studies of total HIV DNA [37, 34, 45, 73]. Therefore, in studies of ART treated study participants, many observed singleton sequences may actually be members of large proliferative clones [59].

### 2.8 Predictions are in keeping with recent study

In a recent study of untreated individuals, clones were not observed in 3 participants sampled during Fiebig III-IV stages of infection (days 22-27); but 4 participants sampled during Fiebig stage IV-V (∼day 30-70 post infection) had at least one detectable HIV DNA clone based on chromosomal integration site analysis; [17]. The number of HIV DNA integration sites sampled in the Fiebig IV-V participants varied from 48 - 709. The mean percentage of clones among all sequences in the Fiebig stage V participants was extremely low (0.72%).

Results from this study are in keeping with our model’s predictions (figure 7). We performed sampling of HIV DNA sequences from model-generated data to identify sequence sample size required for detection of clones at different times post-infection (corresponding to distinct Fiebig stages). The fraction of the sample identified as clonal is shown in figure 7.

**Figure 7:**
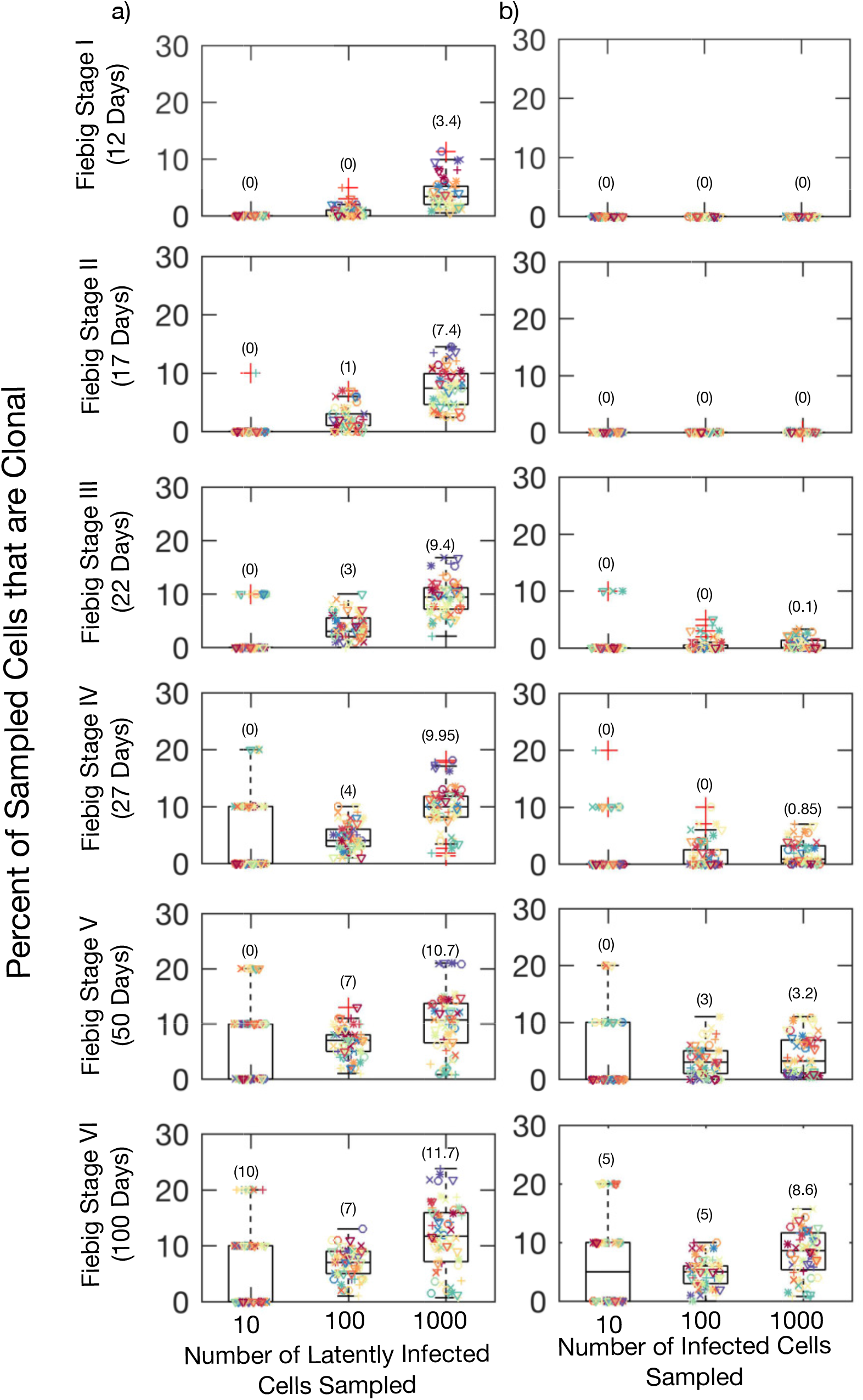
Increased likelihood of detection of proliferative clones within the reservoir at later sampling times and greater HIV DNA sequence sample size from stochastic model simulations. Sampling from: a) Latently Infected Cells only, b) All Infected Cells. Each colour represents simulation results for an individual as in figure 3 and each symbol represents one of five simulation results. The number

If technology existed to sample only HIV DNA sequences from the latently infected reservoir, then sampling of 1000 sequences would reliably detect clones by Fiebig I while sampling of 100 sequences would usually detect clones by Fiebig III (figure 7, left column). However, with sampling of all HIV DNA sequences (figure 7, right column), clones would almost never be evident during Fiebig stages I-III. A sequence sample size of 1000 would sometimes allow detection of a low percentage of clones during Fiebig IV as in [17] and usually allow their detection by Fiebig stage V. A sequence sample size of 100 would reliably detect clones only after 2 months of untreated infection (Fiebig V) (figure 7).

These results are dependent on the mean and variance of turnover rates. Lower mean (figure S5a) and variance (figure S5b) of latent cell proliferation rates results in a lower probability of detection of clones 50 days after infection. Nevertheless, if any latent reservoir proliferation occurs our model predicts that proliferative clones should be possible from Fiebig Stage III of active infection given a sequence sample size of 100 HIV DNA sequences.

### 2.9 Homeostatic proliferation establishes the clonal distribution of the latent reservoir within 2-3 weeks of primary infection

We demonstrated previously that the HIV reservoir has a fixed clonal structure consisting of a small number of large clones and a large number of smaller clones. The rank of each clone according to its size can be used to roughly estimate its size based on a power law relationship with a slope that approximates one [59].

In figure 8 we compare the clonal distribution of the latent reservoir within a single individual at different Fiebig stages. Clonal structure within the simulated reservoir is evident by Fiebig stage II (∼day 17), immediately after recovery from CD4+ T cell nadir (figure 8, orange). The uneven homeostatic proliferation of individual HIV DNA clones in the reservoir is sufficient to explain HIV reservoir structure. As infection progresses, the largest clones comprises a greater proportion of the reservoir (figure 8 a, left). This remains true even if we consider the indistinguishable HIV sequences arising from actively infected cells. A long tail of singleton sequences, clones containing only one member *L*_*n*_ = 1, is observed at all Fiebig stages during untreated infection (figure 8 a, right). The rank-order distributions are also somewhat steeper at later Fiebig stages as the richness of the reservoir decreases (figure 8a).

**Figure 8:**
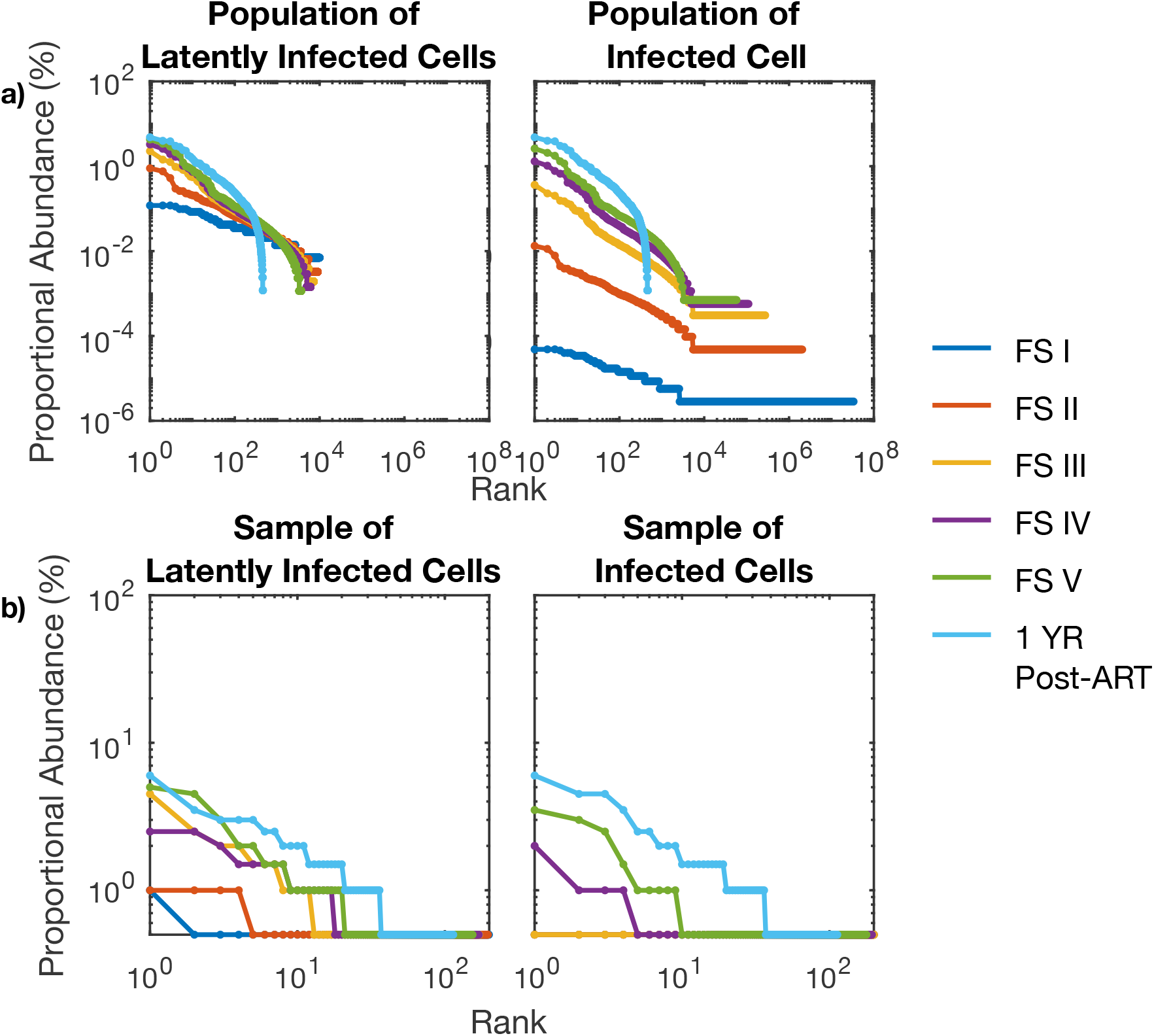
Rank abundance curves from stochastic model simulations for a single participant during sequential stages of early HIV infection. a) Rank abundance curves of all HIV DNA sequences in latently infected cells only (left) and all infected cells (right). b) Rank abundance curves of 200 randomly sampled HIV DNA sequences in latently infected cells only (left) and all infected cells (right).

When we simulated the reservoir after one year of ART started at Fiebig Stage V (100 days post-infection), the clonal distribution of the simulated reservoir after 1 year of effective ART treatment is similar to that of the point at which ART was initiated, though the singleton tail is effectively eliminated. Notably, the rank-order distribution is equivalent between all infected cells and latently infected cells after a year of ART due to the elimination of actively infected cells (figure 8a light-blue). Next we carried out an equivalent analysis assuming clinical sampling of 200 HIV DNA sequences (figure 8 b). Only at Fiebig stages IV and V were multiple clones detected with sampling of sequences from all infected cells, despite our prediction that the clonal structure is established 2 weeks earlier. The distributions we observed after one-year of ART resembled clonal distributions observed during ART in study participants [59], exhibiting a tail of observed singletons which are in fact likely to be members of larger clones.

Varying the mean proliferation rate of clones does not impact the rank-order slope (figure S6a) or the observed distribution of clone frequency assuming known sampling limitations (figure S6b). With no proliferation, our model projects that the reservoir would be composed entirely of singletons and the clonal distribution would not be distinct from a uniform distribution (Kolmogorov-Smirnoff test, p¿0.05). The clonal distribution contains a slightly greater proportion of large clones at a higher proliferation rate relative to a lower proliferation rate (Kolmogorov-Smirnoff test, p¡0.05). This distinction is lost with a sequence sample size of 200 (figure S6b).

The clonal distribution becomes steeper with increasing variance of the proliferation rates due to steeper rank abundance curves at higher variances (figure S6c) (Kolmogorov-Smirnoff test, p=0.05). This distinction is maintained even with a sequence sample size of 200 (figure S6d).

## 3 Discussion

The primary obstacle to eradication of HIV is a reservoir of latently infected resting memory CD4+ T cells [70, 57]. Our mathematical model predicts that the HIV reservoir establishes a steady population during primary infection due to rapid, blind and uneven homeostatic CD4+ T cell proliferation. At the core of our assumptions is that HIV infected reservoir cells behave according to rules governing overall T cell dynamics during recovery from lymphopaenia [74].

Our model generates realistic trajectories for HIV viral load, CD4+ T cell contraction and re-expansion, and CD8+ T cell count expansion during primary infection. The model also recapitulates the reservoir’s early formation, its ultimate size and its clonal distribution. Importantly, our model’s results are consistent with recent data showing that HIV DNA clones become detectable in approximately 50 % HIV infected individuals 1-2 months after primary infection[17].

Sampling the reservoir during early infection is difficult for two reasons. First, most observed HIV DNA sequences are from actively infected cells that have a much shorter lifespan than reservoir cells, harbour truly unique HIV DNA sequences and cannot be discriminated experimentally from reservoir-derived sequences [10]. Second, current studies dramatically under sample the reservoir: during prolonged ART, sequences which are only observed once are likely to be members of large clones rather than truly unique[59]. Despite these issues, our model makes the prediction that repeated HIV DNA integration sites can sometimes be detected within the first 1-2 months of HIV infection, consistent with the results in [17]. While doublets and triplets, clones with two or three members respectively, are only sporadically observed at these timepoints even with large sample sizes, we predict that by this timepoint, there is already a fully formed, mature HIV DNA reservoir with an organised clonal structure. In fact, the doublets and triplets are likely to represent extremely large clones whose size is obscured by under sampling, consisting of more than a million cells each. This signifies that certain clones may undergo roughly twenty divisions during the 2-3 weeks recovery period from CD4+ lymphopaenia.

While observed heterogeneity in clone size may be linked mechanistically with known properties of homeostatic proliferation, we cannot rule out the possibility that the largest clones may be generated due to recognition of a cognate antigen presented by HIV-infected cells or by cells infected with other ubiquitous chronic viruses such as cytomegalovirus or Ebstein-Barr Virus.

Our conclusions demonstrate that cure interventions, even those given during primary infection, face an enormous challenge. Even limited seeding of the reservoir will be followed by a period of homeostatic proliferation during recovery from CD4+ nadir that will expand the size of the reservoir ∼10-fold. Lymphocyte anti-proliferative agents have been proposed as a means to reduce reservoir size during chronic ART [60]. While this approach might be considered in an adjunctive fashion during primary infection, the immunosuppressive features of these drugs raise concerns that they may hamper the long-term cytolytic immune response to HIV. Based on comparable experience following stem cell transplantation [60], it is also questionable whether these agents could sufficiently suppress the powerful homeostatic forces underlying recovery from lymphopaenia.

Our model does reinforce the utility of ART during the first week of acute HIV in limiting reservoir volume [5]. Presumably, this intervention would limit the extent of reservoir seeding, and also limit the extent of CD4+ T cell nadir decline, thereby decreasing the extent of homeostatic proliferation required to reconstitute the overall CD4+ T cell population.

Several important limitations of our study are worth noting. First, while we establish that recovery from CD4+ lymphopaenia and uneven homeostatic proliferation are sufficient to generate the observed clonal structure of the HIV reservoir and argue that the reservoir is formed rapidly during primary infection, key reservoir-sustaining events continue to occur through all subsequent stages of infection. HIV DNA clones wax and wane in size during chronic ART, which suggests that substantial dynamics exists within the reservoir years after recovery from lymphopaenia [74]. These clonal expansions and contraction may relate to recognition of cognate antigen by certain clones rather than a homeostatic process [74].

Moreover, a majority of reservoir sequences observed during prolonged ART arise from timepoints immediately before ART initiation, suggesting that the biologic events which generate reservoir cells continue after primary infection [12, 1]. ART initiation is also followed by a surge in levels of activated, proliferating CD4+ T cells [38]. This process could also further expand a pre-existing reservoir. Neither of these processes is mutually exclusive of, homeostatic proliferation as a driver of reservoir formation during primary HIV infection, and the latter has substantial parallels.

Second, in this study, we focus on total HIV DNA, but only a small fraction of these sequences is replication-competent [13]. As a result, the clones that have been discovered during early HIV infection are probably not replication competent. Nevertheless, viral rebound in patients who initiate ART after only days of infection proves that the replication-competent reservoir is created extremely early during primary HIV [22, 41, 28]. Because replication competent HIV periodically reactivates and is targeted by the immune system, and because viral activation is unpredictably coupled with cellular proliferation [51], it remains unclear whether all conclusions drawn from modelling total HIV DNA during primary infection can be extrapolated to the replication competent reservoir.

Finally, our most parsimonious model does not include an immune response to the virus during primary infection. Our objective was not to identify different drivers of viral load set point or CD4+ T cell nadir, and thus we opted for a simpler model. In our model, the CD4+ and CD8+ T cell equation does not incorporate the possibility that as many as 5-20% of these cells may be reacting to HIV-infected cells and therefore have proliferation governed by HIV antigen abundance rather than homeostasis or expansion due to exposure to other non-HIV antigens [52]. However, we performed deterministic simulations including CD8+ T cell activation which suggest that this effect is unlikely to negate the critical role of homeostatic proliferation in rapidly restoring lymphocyte count following depletion.

In summary, we developed the hypothesis that blind, uneven homeostatic T cell recovery from CD4+ lymphopaenia drives the bulk of early HIV reservoir generation. Future curative therapies will need to address the mechanisms that drive the expansion behaviours of individual T cell clones.

## Data Availability

No data associated.

## Notes

### Competing Interest Statement

The authors have declared no competing interest.

### Funding Statement

This work was supported in part by the NIH Martin Delaney Fund and amfA.

